# Placental Abnormalities of Malperfusion, Inflammation, and Meconium Precede Persistent Pulmonary Hypertension of the Newborn

**DOI:** 10.1101/2025.09.05.25335190

**Authors:** Stephanie M Tsoi, Cynthia Gasper, Emin Maltepe, Megan A Chidboy, Nida Ozarslan, Christine A Blauvelt, Sirirak Buarpung, Shannon Cheung, Martina Steurer, Roberta L Keller, Jeffrey Fineman, Stephanie L Gaw

## Abstract

**Background:** Persistent pulmonary hypertension of the newborn (PPHN) is a cause of neonatal hypoxic respiratory failure due to the failed transition of the pulmonary vasculature after birth. Mechanisms of disease are unknown, but we hypothesize they are directly related to insults in the intrauterine environment. The objective was to describe and compare placentas of PPHN infants to understand significant preceding factors from the maternal-fetal environment.

**Methods:** We conducted a case-control study of mother–infant dyads ≥35 weeks gestation who delivered at a tertiary care center between 2020–2025. Cases were infants diagnosed with PPHN and treated with inhaled nitric oxide; controls were infants without congenital anomalies. Placentas underwent blinded histopathologic review using standardized criteria.

**Results:** 106 placentas were analyzed (53 PPHN, 53 controls). Placental lesions were significantly more common in PPHN, including maternal vascular malperfusion (30.2% vs 9.4%, p<0.01), fetal vascular malperfusion (34.0% vs 17.0%, p=0.05), placental inflammation (66.0% vs 37.7%, p<0.01), meconium (43.4% vs 15.2%, p<0.01), and chorangiosis (7.6% vs 0%, p=0.04).

**Conclusion:** PPHN placentas demonstrate lesions of malperfusion, inflammation, and chronic meconium exposure, suggesting a complex interplay between intrauterine hypoxia and inflammation as a mechanism for the abnormal pulmonary vascular reactivity see in PPHN.

## Introduction

Persistent pulmonary hypertension of the newborn (PPHN) is due to the failure of the pulmonary vascular resistance to fall during transition after birth. It presents as hypoxic respiratory failure in infants within the first hours or days of life. The incidence of PPHN in late preterm and term infants is 0.18% of live births, with a 1-year mortality rate of 7.6%^1^. Multiple etiologic mechanisms may lead to PPHN, including the more common, typical perinatal causes (e.g. sepsis and meconium aspiration syndrome) and the rarer fetal developmental causes (e.g. neonatal interstitial lung disease and lung hypoplasia syndromes such as congenital diaphragmatic hernia)^2^.

The underlying mechanisms of PPHN are poorly understood. While PPHN from fetal developmental causes is often secondary to underdevelopment of the lung and thus its dependent vasculature^3^, typical PPHN is less well defined. Typical PPHN occurs in lungs that have developed normally throughout gestation, suggesting a maladaptation of pulmonary vasculature. Knowledge from animal and human models of pulmonary arterial hypertension suggest that hypoxia, shear stress, inflammation, and genetic influences may contribute to dysfunctional pulmonary vascular reactivity; this may be relevant to PPHN if these mechanisms are present prenatally^4,5^.

Given the presentation of PPHN soon after birth, we hypothesize that the intrauterine environment has a crucial role in pulmonary vascular development, function, and transition. The placenta is the most important organ during pregnancy, responsible for in utero gas and nutrient exchange. Highly vascularized, it serves as the interface between mother and fetus. The objective of this study was to investigate the placental factors that precede PPHN to elucidate potential mechanisms of disease.

## Materials and Methods

This is a case-control study of mother-infant dyads who delivered between January 2020 to May 2025 at the University of California, San Francisco Birth Center. The Birth Center has a community-based delivery service as well as a referral-based delivery service as a fetal diagnostic and interventional center with coordinated delivery and neonatal care for fetuses with anomalies and other conditions requiring specialized perinatal care. Cases were identified if an infant met all of the following criteria: had hypoxemic respiratory failure, diagnosed with PPHN, received inhaled nitric oxide treatment for at least 4 hours, and had placental pathology sent at the time of birth. Controls were randomly chosen amongst mothers who had consented to donate their placenta for research and given birth during the study time frame in the University of California, San Francisco Birth Center. Starting with the most recent samples, charts were queried to identify infants without major congenital anomalies until an equal number of controls to cases were chosen. The inclusion criteria for both cases and controls was gestational age greater than or equal to 35 weeks. This study was approved by the University of California, San Francisco institutional review board: PPHN cases were approved for a waiver of consent, given the retrospective study design. Controls provided written informed consent for sample collection and review of medical records (IRB # 21-33576,#20-32077).

### Demographic and Clinical Data

Electronic medical records from mother-infant dyads were reviewed to obtain information on demographics, maternal co-morbidities, pregnancy complications, delivery information, and neonatal outcomes. Infant race and ethnicity were self-reported by the primary caregiver. Fetal growth restriction (FGR) was defined as having an abdominal circumference or estimated fetal weight below the tenth percentile for gestational age. Small for gestational age (SGA) was defined as having a birth weight below the tenth percentile for gestational age. Advanced maternal age (AMA) was defined as having a maternal age greater than or equal to 35 years. Cases of PPHN were further classified into two groups based on their etiology of disease: 1) typical PPHN (from perinatal causes such as meconium aspiration syndrome or sepsis, and idiopathic cases) and 2) developmental PPHN (from fetal developmental causes like interstitial lung disease, pulmonary hypoplasia, or genetic syndromes)^2^.

### Placental Collection and Processing

At the time of delivery, placentas were sent for gross and microscopic histopathologic examination or collected for research and fixed in 10% buffered formalin. Two to four full-thickness sections from the chorionic plate to the basal plate were taken of each placenta. Sections underwent routine processing, were paraffin embedded, sectioned at 4 μm, and stained with hematoxylin and eosin. A single subspecialist pathologist (C.G.), who was blinded to clinical data except gestational age, reviewed the entirety of each slide under light microscopy (magnification 4 to 40x), and categorized the pathologic lesions according to the Amsterdam Criteria^6^. Placental pathologic findings of interest included any evidence of maternal vascular malperfusion, fetal vascular malperfusion, and inflammation, as well as findings of meconium, chorangiosis, or intervillous thrombi (Supplemental Table 1).

### Statistical Analysis

Descriptive data were reported as mean with standard deviation or median with IQR, and counts with percentages. The PPHN groups were compared to each other (typical vs developmental) and PPHN cases to controls via t-test for continuous variables and by chi-squared analysis for categorical variables. Univariate logistic regression modeling was used to detect associations between placental findings; results were reported as an odds ratio with a 95% confidence interval. Statistical significance was set at a p-value ≤ 0.05. All analyses were performed using Stata SE (Statacorp 2021. Stata Statistical Software: Release 17).

## Results

A total of 106 placentas were included in the analysis: 53 cases with PPHN, 22.6% (n=12) with typical PPHN and 77.4% (n=41) with developmental PPHN, and 53 controls. Maternal gravidity, infant gestational age and infant sex were similar across groups. More mothers in the PPHN group compared to controls had gestational hypertension (32.1% vs 15.1%, p = 0.04), obesity (37.7% vs 12.0%, p < 0.01), and fetal growth restriction (20.8% vs 6.0%, p = 0.03). Infants with PPHN had higher rates of neonatal intensive care unit (NICU) admission (100% vs 22.6%, p < 0.01) and increased mortality (17.0% vs 0%, p < 0.01). (Table 1)

**Table 1.**
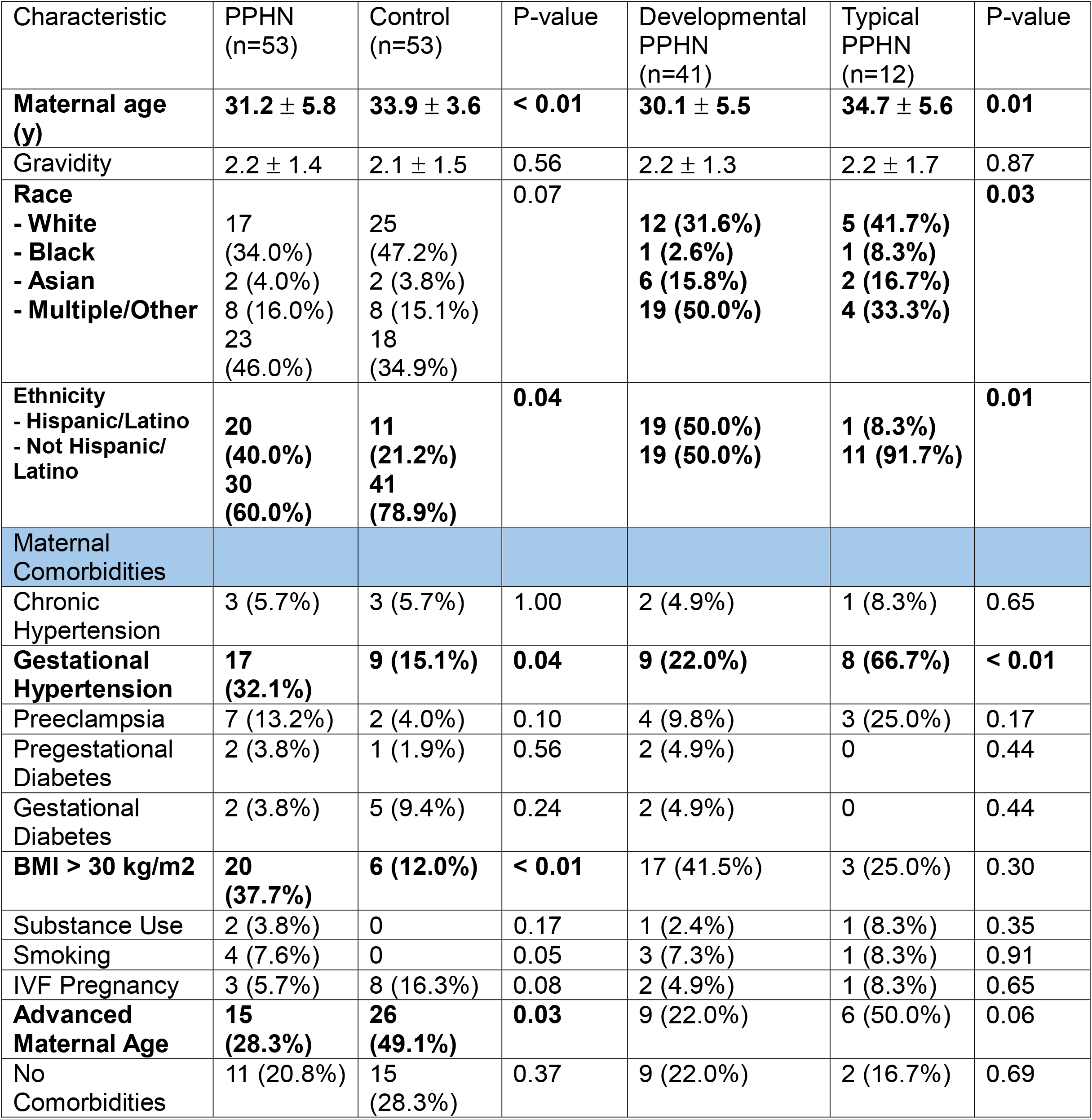
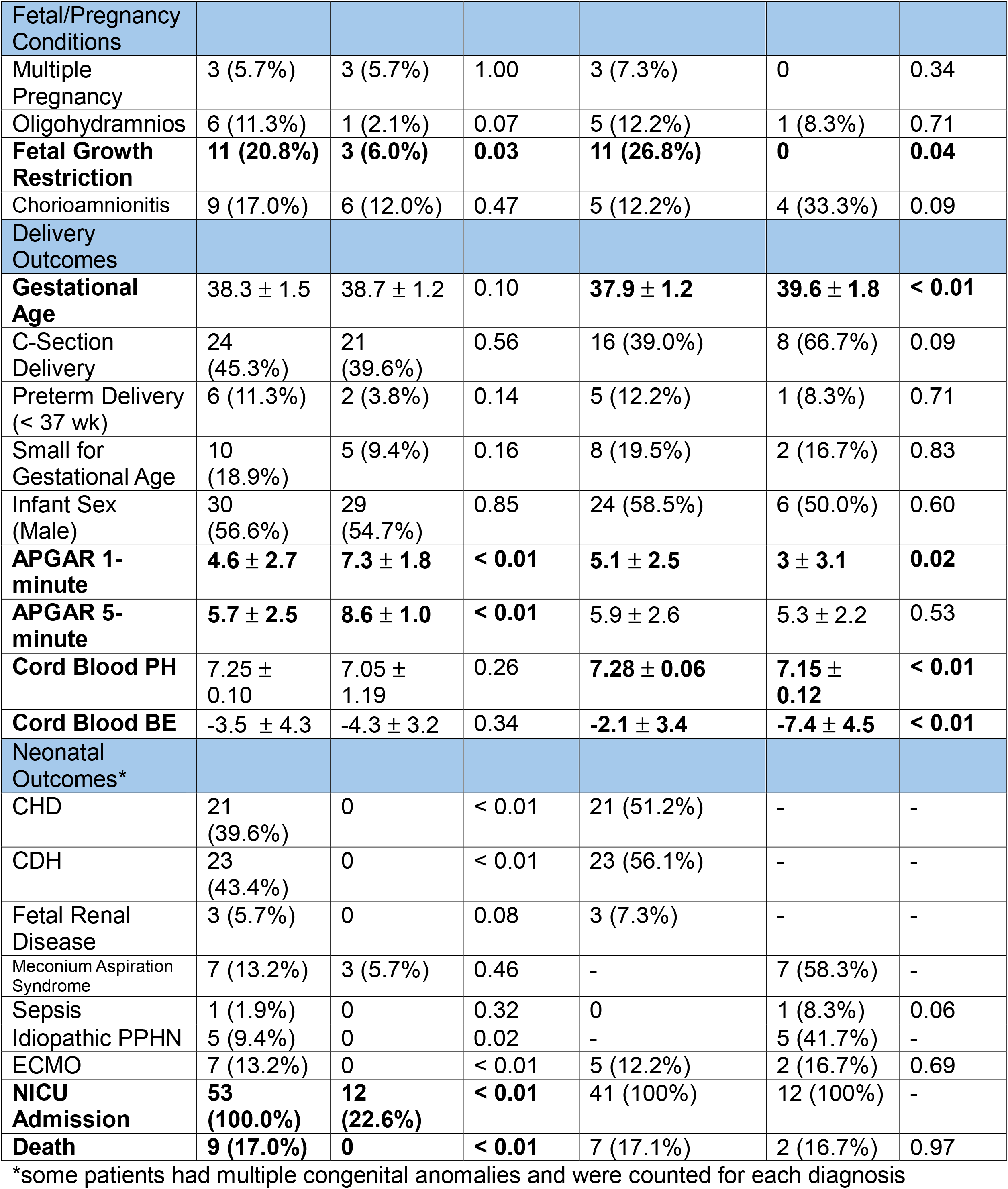
Demographic and clinical characteristics of the patient cohort, separated by cases and controls, and by etiology of PPHN (developmental and typical).Abbreviations: BMI: body mass index, IVF: in-vitro fertilization, APGAR: appearance, pulse, grimace, activity, respiration, BE: base excess, CHD: congenital heart disease, CDH: congenital diaphragmatic hernia, ECMO: extracorporeal membranous oxygenation, NICU: neonatal intensive care unit.

Placental lesions occurred more frequently in PPHN placentas than in controls (92.4% v 60.4%, p < 0.01). The placental pathology associated with PPHN included lesions of maternal vascular malperfusion, fetal vascular malperfusion, placental inflammation (both maternal and fetal inflammatory response), meconium, and chorangiosis (Table 2a). The most common lesion of fetal vascular malperfusion was avascular villi (22.6% of PPHN placentas versus 5.7% of control placentas, p = 0.01) (Figure 1.C-D). Further, chorangiosis, a finding of increased capillary growth in the terminal chorionic villi secondary to hypoxia, was only found in PPHN placentas (7.6%, p = 0.04) (Figure 1.H). 52.8% of PPHN placentas had evidence of a maternal inflammatory response (p = 0.03) and 43.4% had evidence of a fetal inflammatory response (p < 0.01) (Figure 1.E-F). Meconium was seen in the chorion or inside chorionic macrophages, a pattern suggestive of subacute or chronic meconium exposure, in 43.4% of PPHN placentas and 15.1% of controls (p < 0.01) (Table 2a, Figure 1.E, 1.G).

**Table 2.** Comparison of histologic placental pathology by Amsterdam Criteria across A) PPHN placentas and control placentas, B) developmental PPHN and typical PPHN, and C) developmental PPHN and controls, as well as typical PPHN and controls.

| A. | PPHN<br>(n=53) | Control<br>(n=53) | P-value |
| --- | --- | --- | --- |
| Fetal Vascular Malperfusion | **16 (30.2%)** | **5 (9.4%)** | **< 0.01** |
| Maternal Vascular Malperfusion | **18 (34.0%)** | **9 (17.0%)** | **0.05** |
| Placental Inflammation | **35 (66.0%)** | **20 (37.7%)** | **< 0.01** |
| Maternal Inflammatory Response | **28 (52.8%)** | **17 (32.1%)** | **0.03** |
| Fetal Inflammatory Response | **23 (43.4%)** | **3 (5.7%)** | **< 0.01** |
| Other Findings |  |  |  |
| - Meconium | **23 (43.4%)** | **8 (15.1%)** | **< 0.01** |
| - Chorangiosis | **4 (7.6%)** | **0** | **0.04** |
| - Intervillous thrombi | 4 (7.6%) | 2 (3.8%) | 0.40 |
| No Lesions | **4 (7.6%)** | **21 (39.6%)** | **< 0.01** |

| B. | Developmental<br>PPHN<br>(n=41) | Typical<br>PPHN<br>(n=12) | P-value |
| --- | --- | --- | --- |
| Fetal Vascular Malperfusion | 12 (29.3%) | 4 (33.3%) | 0.79 |
| Maternal Vascular Malperfusion | 14 (34.2%) | 4 (33.3%) | 0.96 |
| Placental Inflammation | 26 (63.4%) | 9 (75.0%) | 0.47 |
| Maternal Inflammatory Response | 19 (46.3%) | 9 (75.0%) | 0.08 |
| Fetal Inflammatory Response | 15 (36.6%) | 8 (66.7%) | 0.06 |
| Other Findings |  |  |  |
| - Meconium | **14 (34.2%)** | **9 (75.0%)** | **0.01** |
| - Chorangiosis | 2 (4.9%) | 2 (16.7%) | 0.17 |
| - Intervillous thrombi | 3 (7.3%) | 1 (8.3) | 0.91 |
| No Lesions | 4 (9.8%) | 0 | 0.26 |

| C. | Developmental<br>PPHN<br>(n=41) | Control<br>(n=53) | P-value | Typical<br>PPHN<br>(n=12) | Control<br>(n=53) | P-value |
| --- | --- | --- | --- | --- | --- | --- |
| Fetal Vascular Malperfusion | **12 (29.3%)** | **5 (9.4%)** | **0.01** | **4 (33.3%)** | **5 (9.4%)** | **0.03** |
| Maternal Vascular Malperfusion | 14 (34.2%) | 9 (17.0%) | 0.06 | 4 (33.3%) | 9 (17.0%) | 0.20 |
| Placental Inflammation | **26 (63.4%)** | **20 (37.7%)** | **0.01** | **9 (75.0%)** | **20 (37.7%)** | **0.02** |
| Maternal Inflammatory Response | 19 (46.3%) | 17 (32.1%) | 0.16 | **9 (75.0%)** | **17 (32.1%)** | **< 0.01** |
| Fetal Inflammatory Response | **15 (36.6%)** | **3 (5.7%)** | **< 0.01** | **8 (66.7%)** | **3 (5.7%)** | **< 0.01** |
| Other Findings |  |  |  |  |  |  |
| - Meconium | **14 (34.2%)** | **8 (15.1%)** | **0.03** | **9 (75.0%)** | **8 (15.1%)** | **< 0.01** |
| - Chorangiosis | 2 (4.9%) | 0 | 0.10 | **2 (16.7%)** | **0** | **< 0.01** |
| - Intervillous thrombi | 3 (7.3%) | 2 (3.8%) | 0.45 | 1 (8.3) | 2 (3.8%) | 0.50 |
| No Lesions | **4 (9.8%)** | **21 (39.6%)** | **< 0.01** | **0** | **21 (39.6%)** | **< 0.01** |

**Figure 1.**
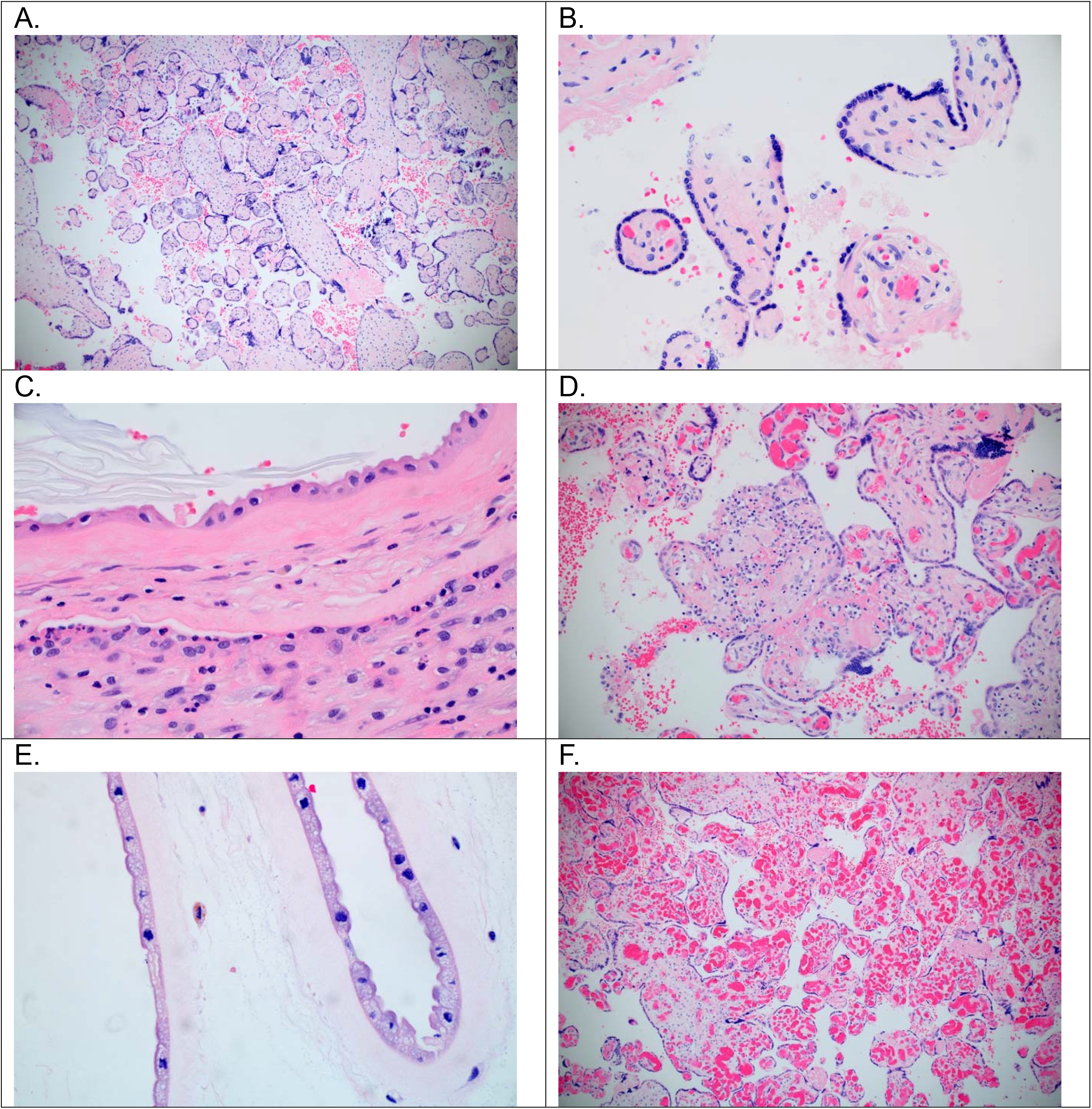
H&E stained slides of placental tissue: A) fetal vascular malperfusion at 10x, B) fetal vascular malperfusion at 40x, C) meconium and acute chorioamnionitis at 60x, D) chronic villitis at 20x, E) meconium at 20x, and F) chorangiosis at 20x.

In univariate logistic regression modeling, histopathologic meconium was associated with placental inflammation in the PPHN group (OR 4.16 (95% CI 1.13, 15.2)), but not in the control group (OR 0.99 (95% CI 0.21, 4.67)). Amongst PPHN patients with meconium (n=23), 82.6% (19/23) also had placental inflammation compared to 37.5% (3/8) of control patients. Amongst PPHN patients with placental inflammation (n=35), 54.3% (19/35) also had meconium compared to 15.0% (3/20) of control patients. Meconium did not show an association with fetal vascular malperfusion in either PPHN (OR 0.67 (95% CI 0.23, 2.01)) or control group (OR 0.39 (95% CI 0.04, 3.53)).

A secondary analysis between typical and developmental PPHN showed that the only placental finding that significantly differed between the two PPHN etiologies was meconium, which was more common in typical PPHN (75.0% versus 34.2%, p = 0.01) (Table 2b). In the presence of meconium, the unadjusted odds ratio for PPHN from typical causes was 5.79 (95% CI 1.35, 24.88). Compared to controls, maternal inflammatory response and chorangiosis were significantly more frequent for typical PPHN, but not for developmental PPHN (Table 2c). There were no placentas without lesions in the typical PPHN group. Lastly, when comparing maternal co-morbidities across PPHN etiologies, gestational hypertension was more prevalent amongst typical PPHN (66.7% vs 22.0%, p < 0.01), while fetal growth restriction was more prevalent amongst developmental PPHN (26.8% vs 0%, p = 0.04) (Table 1).

## Discussion

This is the first study that analyzes the placenta to better understand the maternal-fetal environment that precedes a diagnosis of persistent pulmonary hypertension of the newborn. Infants with PPHN have more placental pathological lesions of malperfusion, inflammation, chorangiosis, and meconium than controls. These findings suggest that hypoxia secondary to malperfusion and inflammation precede PPHN and may be contributing mechanisms to the abnormal pulmonary vascular reactivity.

Similar studies using the placenta have been done to investigate the impact of the intrauterine environment on other neonatal lung-associated pulmonary vascular disease, such as bronchopulmonary dysplasia-associated pulmonary hypertension (BPD-PH); prior work has shown that the presence of maternal vascular underperfusion in placentas from extremely premature infants was associated with later development of BPD and findings consistent with severe placental defects, including decreased villous vascularity, were associated with BPD-PH^7,8^. These data suggest a mechanistic role of intrauterine hypoxia-ischemia exposure in lung and vascular development^7^. Our findings in PPHN placentas similarly suggest a higher prevalence of chronic prenatal hypoxia-ischemia, as well as inflammation and associated meconium. This pathology aligns well with known postnatal mechanisms of pulmonary vascular disease and may indicate that inflammation and hypoxia are early insults contributing to the abnormal development and function of the pulmonary vasculature in PPHN infants.

To understand what drives these insults, we described differences across groups in maternal and fetal conditions that may affect the placenta, thus altering the maternal-fetal environment during gestation or peri-delivery. These included maternal co-morbidities (e.g. hypertensive disorders of pregnancy, diabetes, obesity), fetal conditions (e.g. oligohydramnios, multiple gestation, fetal growth restriction), toxic exposures (e.g. maternal smoking or substance use), and perinatal complications (e.g. chorioamnionitis). The prevalence of gestational hypertension was higher in PPHN placentas, specifically typical PPHN. This may be relevant as hypertensive disorders of pregnancy have known effects on the placenta, resulting in placental lesions of maternal vascular malperfusion leading to impaired oxygen exchange and downstream fetal hypoxemia and oxidative stress^9,10^. Additionally, obesity, smoking, and fetal growth restriction were also significantly higher in the PPHN groups. Interestingly, these listed conditions all cause placental lesions related to impaired placental function, and are all known risk factors for PPHN^1,11–14^. Thus, our findings might suggest a mechanism for how these maternal and fetal conditions are associated with PPHN.

Fetal vascular malperfusion and placental inflammation may reflect how intrauterine insults lead to the pulmonary vascular impairments characteristic of PPHN. First, hypoxia induces pulmonary vasoconstriction and causes pulmonary vascular remodeling^5^. It also alters gene expression in pulmonary endothelial and smooth muscle cells involved in vascular growth and function, such as VEGF, 5-HT, and HIF-1 signaling^15^. Chronic in utero hypoxia exposure in fetuses born to high-altitude ovine pregnancies causes neonatal pulmonary hypertension, with increased vascular reactivity and important molecular changes, suggesting a detrimental effect of early hypoxia on the pulmonary vasculature^16^. Our findings of increased fetal vascular malperfusion and chorangiosis in PPHN placentas suggest that these fetuses are similarly exposed to a chronic in utero hypoxia. These results suggest that hypoxia is driving cellular and molecular changes in the developing pulmonary vasculature, a potential mechanism altered pulmonary vascular reactivity in PPHN.

Second, inflammation and circulating immune factors can also cause pulmonary vascular damage, remodeling, and dysfunction^17^. When immune cells, especially macrophages, aggregate in pulmonary arteries, it worsens vascular remodeling by causing maladaptive fibroproliferative changes^18^. Various cytokines and chemokines are increased in patients with pulmonary hypertension, and have been found to interact with key regulators (e.g. prostacyclin) and pathways (e.g. BMPR2 signaling pathway) of normal pulmonary artery adaptation^19^. Given our finding of a significant association between placental inflammation and PPHN, it is possible that these circulating immune factors in the intrauterine environment pass through the placenta to the fetus and are key contributors to the pathophysiology of PPHN.

We also found the chronic presence of meconium to be significantly associated with PPHN, a finding that held across both typical and developmental PPHN. The presence of meconium in the chorion and chorionic macrophages indicates chronic exposure to meconium from the amniotic fluid, rather than acute peripartum exposure that is commonly noted in deliveries that underwent the labor process. In our study, meconium was seen both free-floating and phagocytosed by the chorionic macrophages, and with associated columnar epithelial changes (cellular changes representative of a longer meconium exposure)^20^. Chronic meconium exposure may support an inflammatory mechanism leading to PPHN, as the association between meconium and placental inflammation was only found in the PPHN group. Meconium could be acting as a toxin, eliciting a placental inflammatory response, much like it does in the pulmonary vasculature in meconium aspiration syndrome-induced PPHN^21^. In addition, the fetus may excrete meconium prenatally in response to poor placental gas and nutrient exchange secondary to fetal vascular malperfusion^22^, thus creating an inflammatory state. This would explain why 82.6% of patients with meconium also had placental inflammation, but only 54.3% of patients with placental inflammation had meconium – suggesting that meconium is triggering the inflammation. In addition to driving placental inflammation, chronic meconium exposure may also contribute directly to abnormalities in fetal pulmonary vascular development. In fatal cases of meconium aspiration syndrome, meconium-stained amniotic fluid is thought to directly cause placental vascular lesions and neonatal pulmonary vascular remodeling^23^. It is possible that chronic meconium exposure in utero may impair fetal pulmonary vascular development, leading to the vascular dysfunction of PPHN.

Our secondary analysis comparing developmental versus typical PPHN demonstrates a potential role for the placental to help predict etiology of PPHN. Given that certain forms of developmental PPHN are associated with higher mortality and necessitate more invasive tests (i.e. advanced imaging, genetic testing, or lung biopsy) before a diagnosis, distinguishing these cases from typical PPHN using the placenta may be informative. While PPHN presents in the first hours or day of life, it can take nearly two weeks for the course of typical PPHN to differentiate from that of developmental PPHN^2^. For example, our data suggests that the presence of meconium on placental histopathology would place that infant at higher odds of their PPHN being from typical causes than developmental causes. Future investigations with larger sample sizes could develop a multiparameter score that better predicts typical PPHN based on placental pathology and a history of maternal or fetal conditions (such as a history of gestational hypertension, more common in typical PPHN, or fetal growth restriction, more common in developmental PPHN).

Overall, this study has multiple strengths. We performed standardized placental pathology review for the purposes of research, by a pathologist blinded to the diagnosis of PPHN, reducing observer bias. We reduced selection bias by having equal case and control groups and randomly selecting controls. This distribution of PPHN groups was not representative of the general population. As a referral center for fetal anomalies, we had a relatively large number of developmental PPHN cases, but a weakness, as noted, was the limited number of typical PPHN cases. Additionally, typical PPHN cases may not have had a clear indication for requesting a placental pathologic analysis, and so there may be selection bias in this group. Given the single-center and retrospective nature of our study, future multi-center and prospective studies are needed to validate our findings.

In conclusion, placentas from infants with PPHN are more likely to have histologic lesions, including fetal and maternal vascular malperfusion, inflammation, meconium, and chorangiosis. These placental lesions reinforce that an abnormal maternal-fetal environment marked by hypoxia and inflammation precede PPHN, and may be mechanistically responsible for the dysfunctional pulmonary vasculature. Further work is needed to fully establish this placental-pulmonary axis and its effects beyond the neonatal period.

## Data Availability

All data produced in the present study are available upon reasonable request to the authors

---

**Supplemental Table S1.** Description of placental lesions based on the Amsterdam Criteria

| Category | Example of diagnoses |
| --- | --- |
| Fetal vascular malperfusion | Vascular thrombi<br>Avascular villi<br>Villous stromal-vascular karyorrhexis<br>Intramural fibrin deposition<br>Stem villous obliteration / fibromuscular sclerosis<br>Vascular ectasia |
| Maternal vascular malperfusion | Placental hypoplasia (weight < 10<sup>th</sup> percentile)<br>Placental infarct<br>Accelerated villous maturation<br>Retroplacental hemorrhage<br>Distal villous hypoplasia<br>Decidual arteriopathy<br>Increased syncytial knots |
| Placental Inflammation | <u>Maternal inflammatory response</u><br>Acute subchorionitis<br>Acute chorioamnionitis<br>Chorionic plate inflammation<br><u>Fetal inflammatory response</u><br>Chronic villitis<br>Umbilical phlebitis*<br>Umbilical arteritis*<br>Funisitis*<br>Chronic Deciduitis |
| Other placental finding | Meconium<br>Chorangiosis<br>Intervillous thrombi<br>Hyper/Hypocoiled cord* |

\*only available in cases, controls did not include the umbilical cord

